# Data release and baseline analysis of the largest collection of 6-minute walk tests: the My Heart Counts Cardiovascular Health Study, a fully digital smartphone platform

**DOI:** 10.1101/2024.06.26.24309535

**Authors:** Daniel Seung Kim, Narayan Schuetz, Anders Johnson, Alexander Tolas, Sriya Mantena, Jack W O’Sullivan, Steven G Hershman, Abby C King, Jeffrey W Christle, Marily Oppezzo, Fatima Rodriguez, C. Mikael Mattsson, Matthew T Wheeler, Herman A Taylor, Susan Murphy, Euan A Ashley

**Author notes:** Denotes equal contribution (Daniel Seung Kim, Narayan Schuetz). <u>Corresponding author:</u> Euan A. Ashley, MB, ChB, DPhil Professor of Medicine and Genetics Falk CVRC, 870 Quarry Road, Stanford, CA 94305.

## Abstract

The six-minute walk test (6MWT) is a sub-maximal exercise test used clinically as a measure of functional capacity. With the emergence of advanced sensors, 6MWTs were commonly performed remotely via smartphones and other devices. The My Heart Counts Cardiovascular Health Study is a smartphone application that serves as a digital platform for studies of human cardiovascular health. It has been used to perform 30,475 6MWTs on 25,539 unique participants. Here, we report on the associations of demographic and clinical variables with 6MWT distance at baseline (N=4,576), validating prior associations with male gender, height, and baseline physical activity with 6MWT distance in multivariable regression analysis. We also report associations of 6MWT baseline distance with working status (+26.8m ±5.2m, P<0.001) and feeling depressed (−3.78m, ±0.79m, P<0.001). In a subset of participants who conducted repeat 6MWTs separated by at least 1-week but no greater than 3-months (N=2,269), we found that use of the *My Heart Counts* app resulted in a statistically significant increase in 6MWT distance (+21.5m ±5.72m, *P*<0.001). Finally, we present the public data release of our 30,475 6MWTs and the launch of a webpage-based data viewer of summary-level statistics, to compare the relative health of an individual by their age, gender, and chronic disease status. Given the importance of 6MWTs in assessment of functional capacity, our publicly-available data will serve an important purpose as a healthy and disease-specific reference for investigators worldwide.

## INTRODUCTION

The six-minute walk test (6MWT) is a safe, frequently used objective test of functional exercise capacity in patients^1^. The 6MWT is often used as an alternative to cardiopulmonary exercise testing for assessment of functional exercise capacity due to its low-cost, ease in performing the test, lack of specialized medical equipment, and ability to be performed without the supervision of a healthcare professional^2,3^. During the clinical version of the test, patients are asked to walk as far as possible along a 30m corridor for 6 minutes, with the primary outcome being their 6MWT distance^1^. 6MWT distance traveled is strongly associated with long-term outcomes in both coronary artery disease^4^ and heart failure^5,6^, and is also frequently used to measure clinical evolution of an individual patient with numerous other chronic diseases (e.g., chronic obstructive pulmonary disease, interstitial lung disease, pulmonary arterial hypertension, and neuromuscular diseases)^2^.

With the advancement of smartphone technology to include motion and inertial sensors, 6MWTs began to be conducted remotely and autonomously through smartphone applications^7,8^. Notably, the Core Motion sensor present in Apple-based iPhones has been validated in test-retest technical validation^9^. Using advanced sensors, numerous studies have been conducted, which validated the digital health approach of performing 6MWTs via smartphone applications in diverse patient populations: ranging from healthy participants^7^ to patients with multiple sclerosis^10^, pulmonary arterial hypertension^11^, mitral and aortic valvular disease^12^, among numerous other conditions and diseases^8^.

The *My Heart Counts* Cardiovascular Health Study is a smartphone application, developed in conjunction with Apple as one of the first “ResearchKit” smartphone applications, to serve as a digital platform for studies of human health and disease^13,14^ (a subsequent Android compatible application was later developed in collaboration with Google)^15^. On release in 2015, the *My Heart Counts* application was the fastest recruited medical study in history, with 40,000 participants recruited in the first three weeks^14^. The *My Heart Counts* app and subsequent studies demonstrated the feasibility of digital recruitment and consent (fully automated, without any dedicated study coordinators), and was one of the first studies to use smartphone sensors for quantification of physiologic parameters, including accelerometry for movement and photoplethysmography for heart rate/rhythm assessment^14^. The *My Heart counts* app was later used to conduct fully digital randomized crossover trials^16^, finding that personalized coaching prompts were superior to a more generic, “one-size-fits-all” approach to encouraging physical activity^17^. In addition, we previously publicly released *My Heart Counts* related data from individuals who consented for broad sharing, to advance scientific insights from unique digital health data that remains, to the best of our knowledge, the largest such available dataset^18^.

Given the importance of 6MWTs in objective assessment of functional capacity, we sought to release a data update (N=30,475) that is >6-fold greater than our prior data release (N=4,990)^18^ and will be the largest publicly available dataset of 6MWTs. We provide baseline analyses of the data, recapitulating epidemiologic findings and also finding that symptoms of depression and employment status affect 6MWT distance. Moreover, we demonstrate that My Heart Counts app use resulted in a significant positive change in 6MWT distance. Finally, we report on a webpage-based data browser that will allow for comparison of an individual’s 6MWT distance as compared to age, gender, and disease-status matched comparisons.

## METHODS

### Ethics Statement

Ethical and institutional review board approval for the study was obtained from Stanford University’s Research Compliance Office (IRB approval number 31409).

### Participant onboarding

The *My Heart Counts* smartphone application was first made available in March 2015 through the Apple App Store to residents of the United States, United Kingdom, and Hong Kong. Participants aged 18 years or older and able to read and understand English were eligible to enroll in the *My Heart Counts* Cardiovascular Health Study. Participants then completed an interactive eConsent process, which included animated icons, concise text, and links for more information^19^. After completion of the eConsent process, participants were asked to e-sign an electronically rendered version of the traditional consent form, after which they were emailed a copy of the signed e-consent document. All sensitive login and PHI data were managed by Stormpath, keeping this data separate from other forms of study data. As part of the eConsent process, participants had the option to either share their data narrowly (with Stanford University researchers only) or broadly (with qualified researchers worldwide). All participants had to make an active choice to complete the consent process, as no default choice was selected^20^.

At first login after informed consent, participants were asked to grant the *My Heart Counts* application access to their HealthKit and Core Motion data. HealthKit is a framework to capture, store, and facilitate the sharing of health and physical activity data collected from iPhone (and later, Apple Watch) sensors between smartphone applications (e.g., Fitbit, Nike+ Run Club, Activity). The *My Heart Counts* application passively captured body measurements (height, weight), physical activity data (flights climbed, step count, stand hours, walking and running distance, workouts), sleep data, health results (blood glucose, hemoglobin A1c, and lipid measurements), and vital signs (blood pressure, oxygen saturation) if they were entered into HealthKit by the user. Core Motion is a coprocessor chip of iPhones (iPhone 5S or newer), which integrates a number of sensor signals including a triaxial accelerometry, gyroscope, compass, and barometer, to estimate the presence of movement, distance traveled, and modality of movement (i.e., walking, running, cycling, or driving).

### Six-minute walk test (6MWT) data collection

On the final day of the My Heart Counts Cardiovascular Health Study (8th day after enrollment), participants were asked to complete several questionnaires and complete a self-administered 6MWT. The 6MWT is a phone guided task, which triggers the collection of global positioning system displacement-based distances, pedometer-based distances, pedometer-based step counts, and accelerometer and gyroscope measurements in both raw and processed formats. In a validation set of 26 6MWTs, the mean error was 3.1m^13^. Of note, the *My Heart Counts* based 6MWT did not require frequent turns along a 30m path, as compared with traditional 6MWTs performed in the clinic^2^.

### Statistical Analysis

All analyses were performed using R (https://www.r-project.org). To determine predictors of 6MWT distance traveled, we used a stepwise linear regression model to identify independent predictors in a subset of participants who completed a 6MWT and provided demographic information and completed surveys on self-reported information related to health (N=4,576). Akaike’s information criterion (AIC) was used to examine the fit of each model, beginning with a base model with gender, age in years, height in cm, and body mass index (BMI) in kg/m^2^. Other baseline survey data and disease status variables were then considered and included in the final stepwise regression model if they independently increased the ability of the regression model to predict 6MWT distance traveled. A false discovery rate (FDR) threshold of P<10^-4^ was used to correct for multiple comparisons.

For change in 6MWT distance traveled, only a subset of participants who completed ≥2 6MWTs separated by at least one week but no greater than three months (N=2,269) were considered. As each participant served as their own control (age, gender, height, disease-status matched), a paired T-test was used to compare changes in 6MWT distance traveled. A *P*<0.05 was considered statistically significant.

### Role of the funding source

The funders had no role in study design, data collection, data analysis, data interpretation, or the writing of this manuscript. Apple (Cupertino, CA, USA) and Google (Mountain View, CA, USA) provided support for the initial development and maintenance of the *My Heart Counts* application for iOS and Android, respectively. The corresponding author had full access to the data and the final responsibility to submit for publication.

### Data Availability

All 6MWT data is available through Synapse (https://www.synapse.org). To balance sharing individual-level 6MWT data with privacy protections for participants, we have instituted the following governance structures: researchers interested in accessing this data should register for a Synapse account, have their Synapse User Profile validated by the Synapse Access and Compliance Team, submit an intended data use statement, and agree to data-specific conditions for use. The overarching conditions for use of this data are summarized previously^18^, but require affirmation that researchers will maintain confidentiality and not attempt to contact or identify research participants for any reason, confirm their commitment to the Synapse Awareness and Ethics Pledge, and agree to the guiding principles of responsible research use and data handling.

### Code Availability

Code will be available on request to the corresponding author.

## RESULTS

Summary data from the 30,475 6MWTs can be visualized through a webpage-based point-and-click interface (https://mhc-6mwts.streamlit.app). As this website aims to provide normative value references, one can enter age, gender, height, weight, disease status, and 6MWT total distance and then compare their 6MWT values compared to a matched distribution in our data, which is the largest known collection of 6MWTs.

Individual-level data is available through Synapse, as summarized in the **Methods**. We record the anticipated decrease in average heart rate during the 6MWT with each progressive decade of life (**Figure 1A**). Example individual-level data from paired smartdevice (e.g., Apple Watch) during 6MWTs is presented in **Figure 1B**, with heart rate in the top panel and accelerometry data in the bottom panel. Of note, this individual turned a total of 3 times, as demonstrated by the showing of the accelerometry (purple) data tracing in **Figure 1B**. For 6MWT total distance, a decrease in total distance traveled was observed for each progressive BMI group (**Figure 1C**). Compared to participants without self-reported cardiovascular disease, we note slight decreases in 6MWT total distance in participants with prior self-reported myocardial infarction, heart failure, and stroke, and a larger decrease in 6MWT total distance in a subset of participants with self-reported pulmonary arterial hypertension (**Figure 1D**).

**Figure 1.**
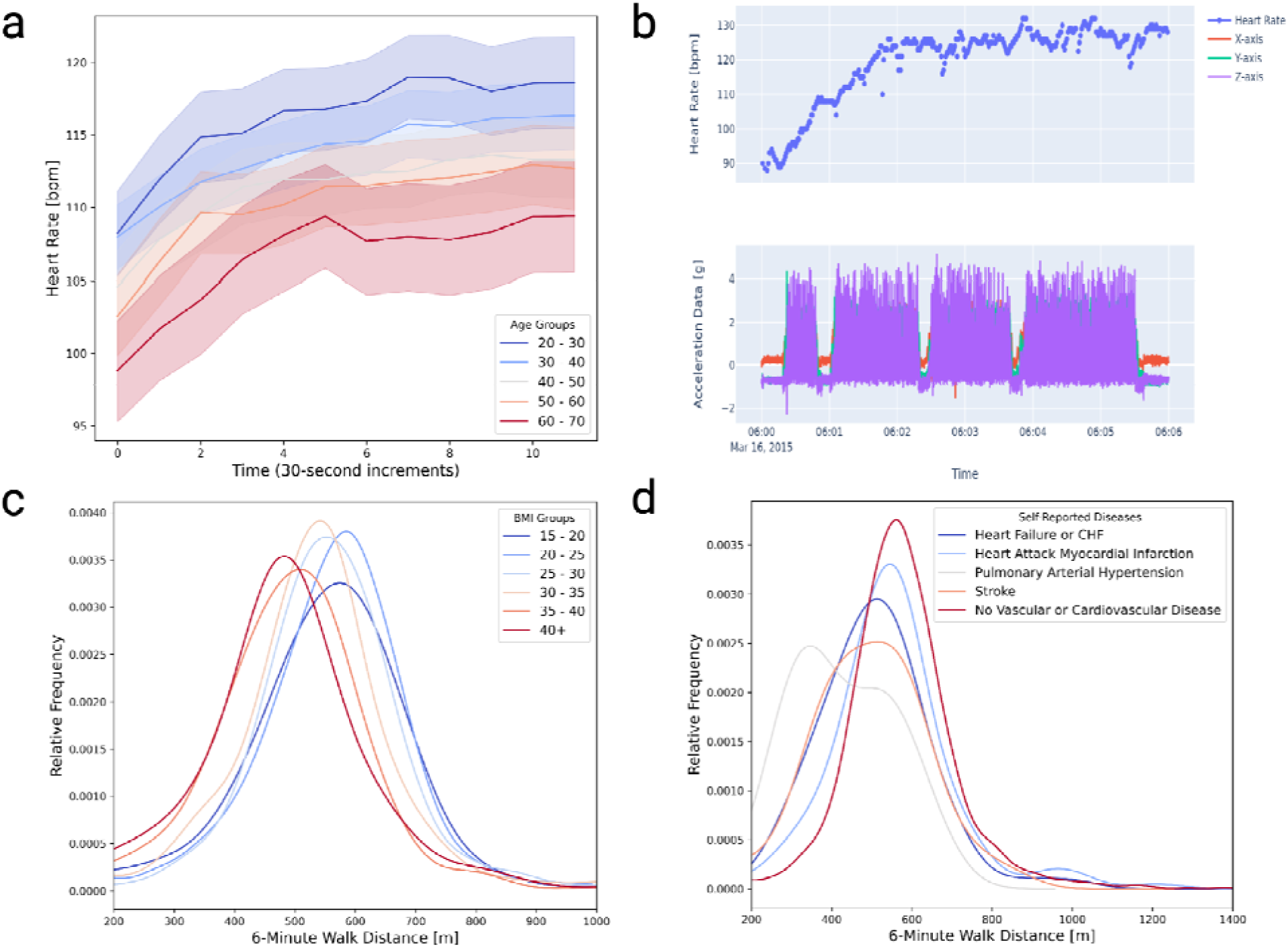
The My Heart Counts Cardiovascular Health Study smartphone application can be used to perform an autonomous, unsupervised, six-minute walk test (6MWT). (a) average heart rate change during a 6MWT by decade of life. (b) example heart rate (top panel) and accelerometry (bottom panel) during a 6MWT from an individual using an Apple Watch, in which a participant stops and turns several times (purple accelerometry tracing). (c) distribution of 6MWT distance by body mass index (BMI) groups. (d) distribution of 6MWT distance by self-reported cardiovascular disease history.

Demographic and clinical characteristics on enrollment to the My Heart Counts Cardiovascular Health Study are presented in **Table 1**. On average, participants were 54.5 years of age and self-identified as white (82%, Black 2.9%, Hispanic/Latinx 4.1%, Asian 8.7%, other 2.2%) men (76%), who had mildly overweight BMI (26.8 kg/m^2^). The majority of participants had either a graduate (38%) or college (34%) degree, with an additional 17.8% having an associate degree, and 7.8% reporting high-school degree as their highest academic level. 78% of participants reported being employed, and with a low overall depression score (2.14, with scaling 1-10). The majority of participants reported meeting American Heart Association guidelines, exercising at moderate intensity at least 3 times weekly. With regard to disease, 17% and 9% reported having heart disease and vascular disease, respectively.

**Table 1.**
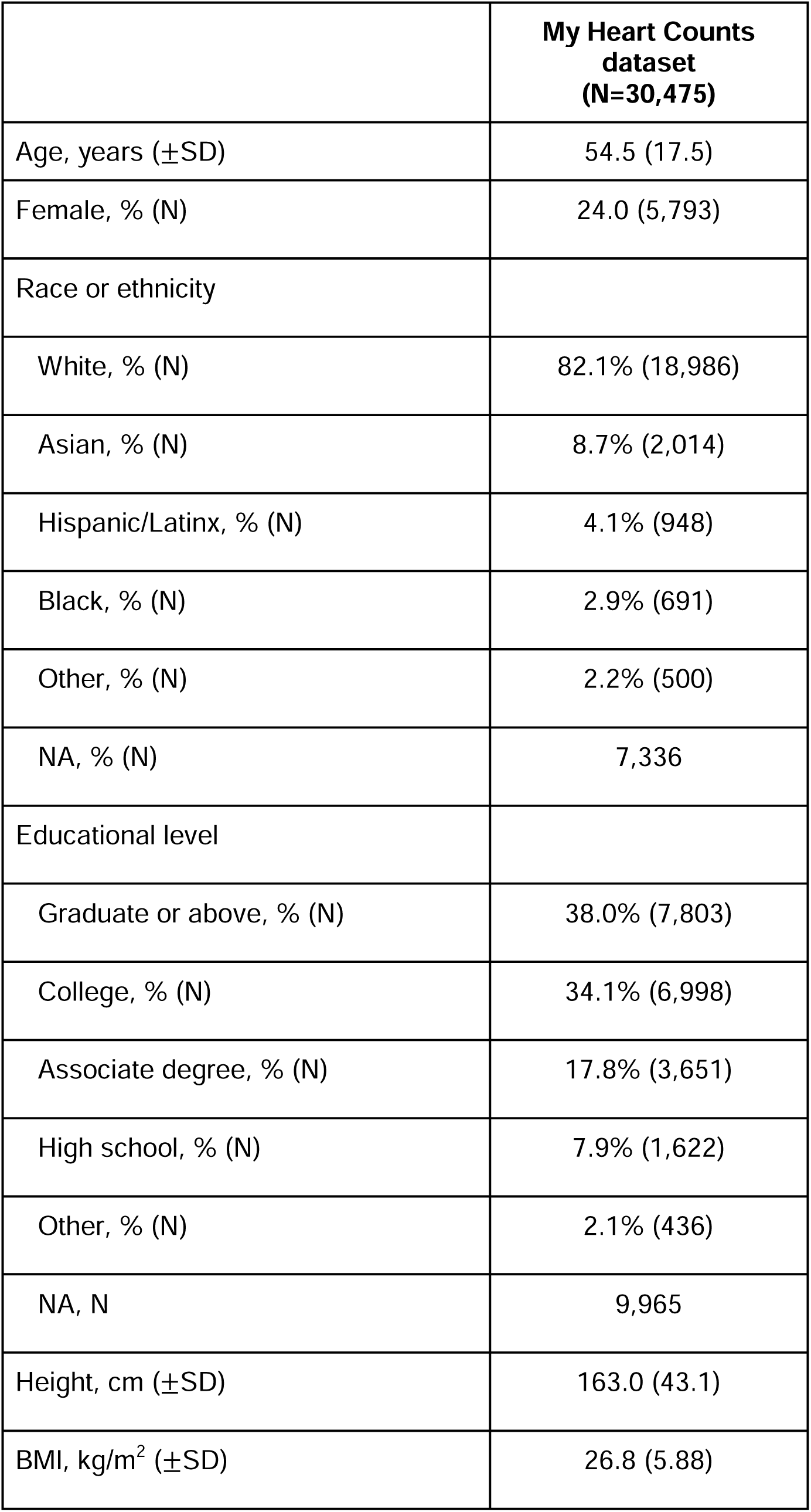
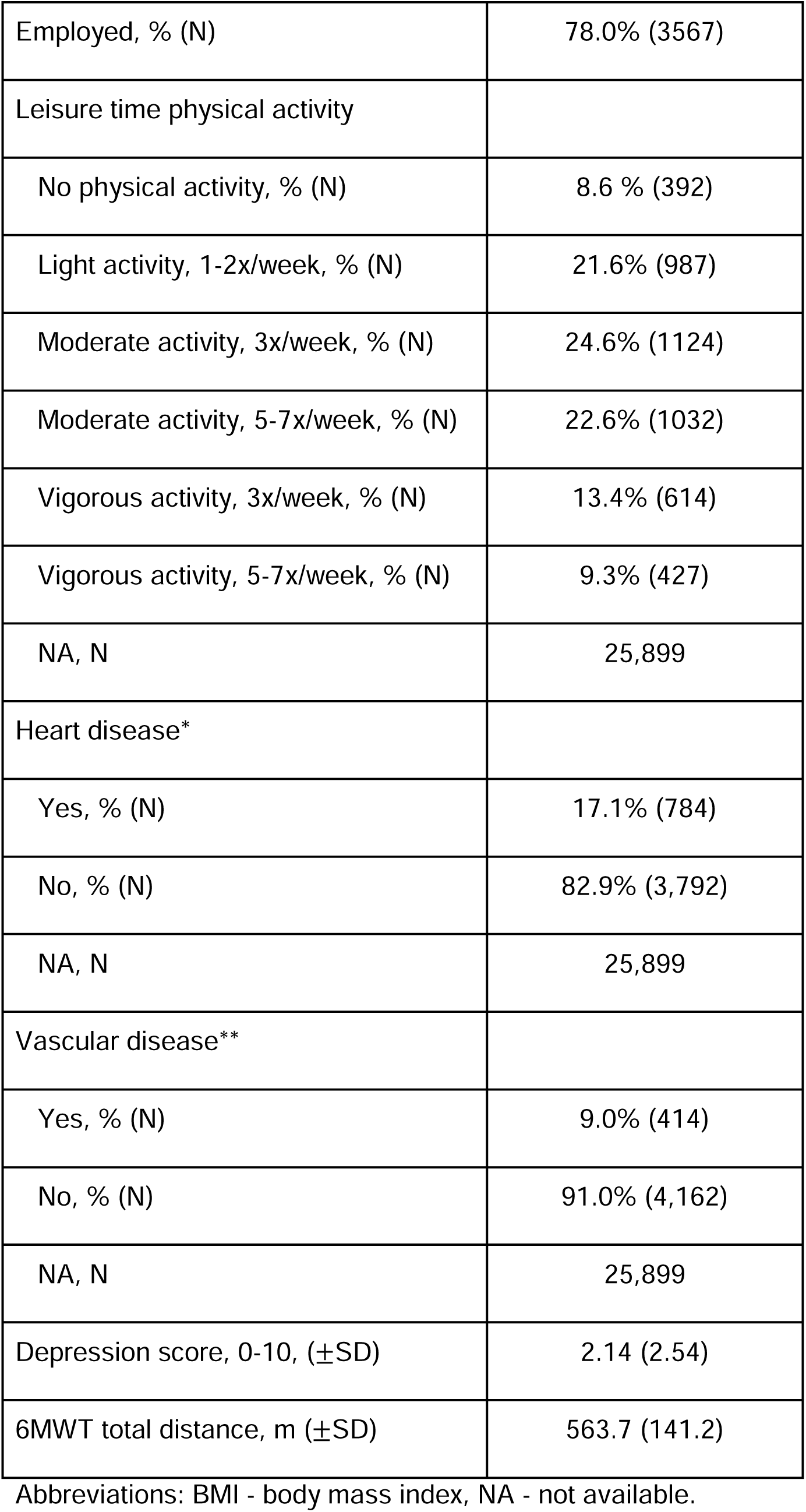
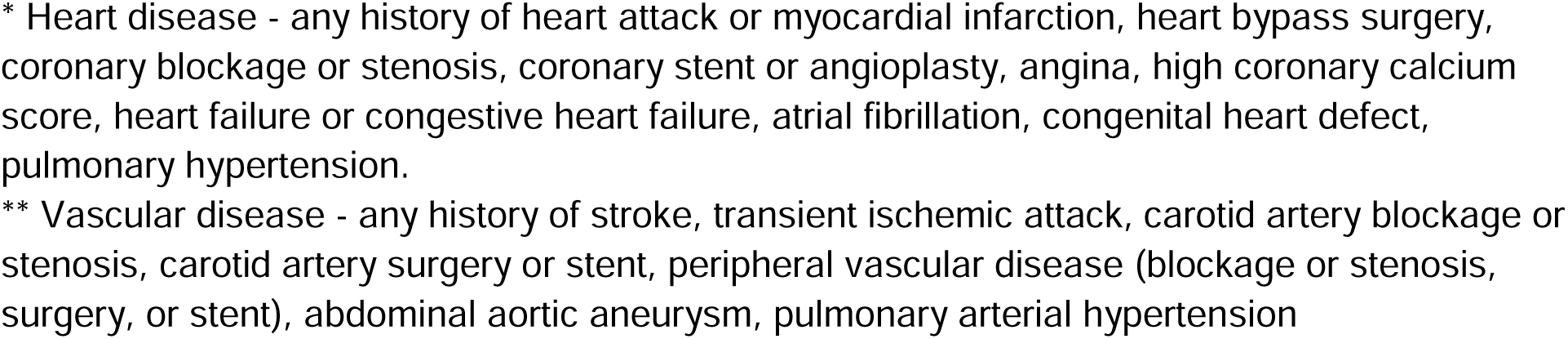
Baseline demographic, clinical, and survey data.

|  | **My Heart Counts**<br>**dataset**<br>**(N=30,475)** |
| --- | --- |
| Age, years (±SD) | 54.5 (17.5) |
| Female, % (N) | 24.0 (5,793) |
| Race or ethnicity |  |
| White, % (N) | 82.1% (18,986) |
| Asian, % (N) | 8.7% (2,014) |
| Hispanic/Latinx, % (N) | 4.1% (948) |
| Black, % (N) | 2.9% (691) |
| Other, % (N) | 2.2% (500) |
| NA, % (N) | 7,336 |
| Educational level |  |
| Graduate or above, % (N) | 38.0% (7,803) |
| College, % (N) | 34.1% (6,998) |
| Associate degree, % (N) | 17.8% (3,651) |
| High school, % (N) | 7.9% (1,622) |
| Other, % (N) | 2.1% (436) |
| NA, N | 9,965 |
| Height, cm (±SD) | 163.0 (43.1) |
| BMI, kg/m<sup>2</sup> (±SD) | 26.8 (5.88) |
| Employed, % (N) | 78.0% (3567) |
| Leisure time physical activity |  |
| No physical activity, % (N) | 8.6 % (392) |
| Light activity, 1-2x/week, % (N) | 21.6% (987) |
| Moderate activity, 3x/week, % (N) | 24.6% (1124) |
| Moderate activity, 5-7x/week, % (N) | 22.6% (1032) |
| Vigorous activity, 3x/week, % (N) | 13.4% (614) |
| Vigorous activity, 5-7x/week, % (N) | 9.3% (427) |
| NA, N | 25,899 |
| Heart disease* |  |
| Yes, % (N) | 17.1% (784) |
| No, % (N) | 82.9% (3,792) |
| NA, N | 25,899 |
| Vascular disease** |  |
| Yes, % (N) | 9.0% (414) |
| No, % (N) | 91.0% (4,162) |
| NA, N | 25,899 |
| Depression score, 0-10, (±SD) | 2.14 (2.54) |
| 6MWT total distance, m (±SD) | 563.7 (141.2) |

Abbreviations: BMI - body mass index, NA - not available.
\* Heart disease - any history of heart attack or myocardial infarction, heart bypass surgery, coronary blockage or stenosis, coronary stent or angioplasty, angina, high coronary calcium score, heart failure or congestive heart failure, atrial fibrillation, congenital heart defect, pulmonary hypertension.
\*\* Vascular disease - any history of stroke, transient ischemic attack, carotid artery blockage or stenosis, carotid artery surgery or stent, peripheral vascular disease (blockage or stenosis, surgery, or stent), abdominal aortic aneurysm, pulmonary arterial hypertension

A base model considering age, gender, height, and BMI was fit for the outcome of 6MWT distance traveled in 4,576 participant’s first 6MWT performed after one-week of onboarding in the My Heart Counts Cardiovascular Health Study. We then examined a best-fit model using stepwise linear regression including the aforementioned variables in the base model, in addition to all demographic, survey, and clinical data presented in **Table 1**. As a result, only variables that improved predictive power of the best-fit model, as determined by Alkaike’s information criterion, were included in the final, full regression model.

In addition to the base model considering age, gender, height, and BMI, three additional survey variables significantly improved model prediction of 6MWT distance traveled: baseline physical activity, employment status, and presence of depressive symptoms (**Table 2**). Of all variables in the final regression model, male gender (□=16.6m, *P*=0.0083), height (□=2.02m per 1 cm, *P*<0.001), baseline physical activity (*P*<0.001), and employment status (□=27.1m, *P*<0.001) were associated with increases in 6MWT distance traveled. Baseline physical activity groups were treated as dummy variables, with the reference group those who reported not doing any leisure time physical activity. We noted an approximately linear, dose-response relationship between baseline physical activity and 6MWT distance, with all groups who did leisure-time physical activity having a significant increase in 6MWT distance traveled (see **Table 2**), ranging from 25.1m (*P*=0.0017) for the light activity group to 86.6m (*P*<0.001) for the vigorous activity >3x per week group. Presence of depressive symptoms on baseline survey on enrollment to the My Heart Counts Cardiovascular Health Study was associated with decreased 6MWT distance traveled when controlling for all other variables (□=-3.65m, *P*<0.001). Of note, age and BMI were not associated with 6MWT distance traveled when adjusting for all other covariates in the final multivariable regression model.

**Table 2.** Results of stepwise linear regression model on 6MWT total distance in 4,576 participants with baseline demographic, survey, and clinical data.

| Variable | Coefficient (± SE) | *P* |
| --- | --- | --- |
| *(Intercept)* | 126.4 (± 45.7) | - |
| Male gender | 16.6 (± 5.94) | 0.0052 |
| Age | -0.092 (± 0.14) | 0.51 |
| Height | 2.02 (± 0.27) | <0.001 |
| BMI | -0.0019 (± 0.0037) | 0.59 |
| Baseline physical activity |  |  |
| No physical activity | - | - |
| Light activity, 1-2x/week | 25.1 (± 7.99) | 0.0017 |
| Moderate activity, 3x/week | 51.9 (± 7.90) | <0.001 |
| Moderate activity 5-7x/week | 68.0 (± 8.11) | <0.001 |
| Vigorous activity, 3x/week | 72.4 (± 8.74) | <0.001 |
| Vigorous activity 5-7x/week | 86.6 (± 9.48) | <0.001 |
| Employed | 27.1 (± 5.21) | <0.001 |
| Depressive symptoms | -3.65 (± 0.79) | <0.001 |

Abbreviations: BMI - body mass index in kg/m<sup>2</sup>.

As the *My Heart Counts* app aimed to improve human health and had a separate trial with coaching e-interventions on the outcome of physical activity, we separately sought to find if *My Heart Counts* application use was associated with changes in 6MWT distance traveled. In a subset of participants who conducted repeat 6MWTs separated by at least 1-week but no greater than 3-months (N=2,269), we found that use of the *My Heart Counts* app resulted in a statistically significant increase in 6MWT distance (+21.5m ± 5.72m, *P*<0.001), as determined by a paired T-test where each participant served as their own control.

## DISCUSSION

Digital health represents a paradigm shift of scale in medical research: while we previously released 4,990 6MWTs to the scientific public^18^, our *My Heart Counts* smartphone application has continued to accrue users and 6MWT data^17^. We now have recorded 30,475 6MWTs, which we release to the scientific community in the largest known collection of such data. Given the clinical utility of 6MWTs^4–6^, our data will serve as an invaluable reference for both healthy and cardiovascular disease specific patients^21–23^.

Compared to other mobile/wearable datasets, the My Heart Counts Cardiovascular Health Study is among the largest publicly available datasets with >50,000 participants. As an example, while the Apple Heart Study has >400,000 participants, individual-level data from HealthKit are not available. All of Us has >15,000^24^, Project Baseline >10,000^25^, and MIPACT >7,000^26^ participants with linked smartwatch data; however of those datasets, only All of Us^24^ and MIPACT are publicly available (MIPACT only as summary data)^26^, and none of these studies measured 6MWTs. Finally, while the UK Biobank has accelerometer data on >100,000 participants, but no heart rate information is linked to the physical activity data^27^. As a result, our data release of 30,475 6MWTs with individual-level data, including heart rate and accelerometry/gyroscopic data, from the My Heart Counts Cardiovascular Health Study represent a unique dataset for researchers to develop training/pre-training algorithms for heart rate and accelerometry/gyroscopic response during 6MWTs.

In addition to individual-level 6MWT data release, we have created a webpage-based data visualization tool (https://mhc-6mwts.streamlit.app). Through our webpage, users can enter in basic demographic information (e.g., age, gender, height, and cardiovascular-related disease status), and then see their predicted 6MWT total distance. If a user has measured their own 6MWT total distance (or their patient’s), they can enter this data in addition to demographic information and see where they fall within the distribution. This data can also be used in the future for creation of updated 6MWT distance reference equations for both the healthy and disease-specific populations^23,28–30^.

In our data, we replicated prior reports of male gender and height being positively associated with 6MWT total distance^31–34^. We do not find previously reported negative associations between age, weight, or BMI and 6MWT distance^30,31,33^. The lack of associations observed in our data may reflect a healthy bias, which tended to be younger and healthy/non-obese individuals, who were early iPhone adopters^13^. The finding of employment status being positively associated with 6MWT total distance, which does not appear to have literature precedence, may similarly reflect the underlying health status of those who are able to work versus those who are unable to work due to significant comorbidities, such as pulmonary arterial hypertension.

We further note a small, but significant dose-dependent effect of depressive symptoms on 6MWT distance in a largely healthy population. This validates prior reports of depression being negatively correlated with 6MWT in patients with heart failure^35,36^ and in the healthy geriatric population^37^. Notably, physical activity is frequently used in the treatment of depression, with numerous trials of exercise training demonstrating improvement in depressive symptoms that are mirrored in 6MWT distance in patients with stroke^38^, fibromyalgia^39^, cancer^40^, atrial fibrillation^41^, and heart failure^42^.

In prior work, we found that personalized coaching interventions delivered as notifications through the *My Heart Counts* app were associated with increased short-term physical activity^17^. Here, we find that use of the *My Heart Counts* app in a subset with repeat 6MWTs had a statistically significant increase in 6MWT total distance by +21.5m. While this observation would support our prior findings that use of our *My Heart Counts* app was associated with increased physical activity^17^ and potentially, cardiorespiratory fitness, interpretation is limited by the learning effect in 6MWTs^2^. The learning effect in 6MWT has most robustly been demonstrated in chronic obstructive pulmonary disease, where patients underwent 6MWTs on two subsequent days and demonstrated an average increase of +27m, possibly due to a lack of familiarity with the test procedures (walking 30m back and forth for 6 minutes)^43^. As a result, two tests are frequently performed clinically, with the first test either discarded or downweighted^2^. To combat this learning effect, we separated the analyzed 6MWTs by at least one week and no greater than three months. As well, there is likely a lesser learning effect in our data, as 6MWTs were encouraged to be outdoors and hence, much more linear and without turns.

Several limitations of our study should be considered. First, our cohort demographics limit the generalizability of our findings. The average subject in our data was a middle-aged white man with a college or greater education, living in the USA, UK, or Hong Kong. As well, this dataset was gathered from iPhone users, who are generally of higher socioeconomic status^44^. In addition, there is a possibility of selection bias, whereby participants who downloaded the *My Heart Counts* app were self-motivated to improve their cardiovascular health and hence do not represent the broader population. Once enrolled in our study, the Hawthorne effect, whereby participants are more active due to knowledge that their activities are being monitored, could subconsciously affect participant’s activity levels^45^. Finally, the My Heart Counts Cardiovascular Health Study was primarily aimed to study short-term interventions and hence cannot extrapolate to longer-term behavioral changes in physical activity^17^.

In summary, we present to the scientific community the release of the largest collection of 6MWTs with individual-level data. We recapitulate known and inferred clinical and epidemiologic associations with 6MWT distance and also concurrently have launched a webpage-based interface for 6MWT reference values. Given the emergence of 6MWTs performed via mobile devices^7,8^ and the importance of 6MWT total distance in prognostication, we anticipate that this collection of data will serve as an invaluable reference for patients and providers alike. These data highlight the promise of scale that is possible through digital health.

## DECLARATIONS OF INTEREST

M.T.W. reports research grant and in kind support from Bristol Myers Squibb and consulting for Leal Therapeutics. E.A.A. reports advisory board fees from Apple and Foresite Labs. E.A.A. and C.M.M. have ownership interest in svexa, outside of the submitted work. E.A.A. has ownership interest in Nuevocor, DeepCell, and Personalis, outside the submitted work. E.A.A. is a board member of AstraZeneca. D.S.K. is supported by the Wu-Tsai Human Performance Alliance as a Clinician-Scientist Fellow, the Stanford Center for Digital Health as a Digital Health Scholar, and NIH 1L30HL170306. The remainder of authors report no potential conflicts of interest.

## Notes

### Author Declarations

Ethical and institutional review board approval for the study was obtained from the Stanford University Research Compliance Office (IRB approval number 31409).

